# Impacts of remdesivir on dynamics and efficacy stratified by the severity of COVID-19: a simulated two-arm controlled study

**DOI:** 10.1101/2020.05.17.20104711

**Authors:** Ting-Yu Lin, Wei-Jung Chang, Chen-Yang Hsu, Chao-Chih Lai, Amy Ming-Fang Yen, Sam Li-Sheng Chen, Hsiu-Hsi Chen

## Abstract

**Background:** The impact of remdesivir on length of stay of hospitalization, high-risk state, and death stratified by the severity of COVID-19 at enrollment is controversial.

**Methods:** We applied a simulated two-arm controlled study design to the data on compassionate use of remdesivir as a secondary analysis. Dynamics of risk states and death from COVID-19 patients defined by the six-point disease severity recommended by the WHO R&D and the time to discharge from hospital were used to evaluate the efficacy of remdesivir treatment compared with standard care.

**Results:** Stratified by the risk state at enrollment, low-risk patients exhibited the highest efficacy of remdesivir in reducing subsequent progression to high-risk state by 67% (relative risk (RR)=0.33,95% CI: 0.30-0.35) and further to death by 55% (RR=0.45, 95%CI: 0.39-0.50). For the medium-risk patients, less but still statistically significant efficacy results were noted in reducing progression to high-risk state by 52% (RR=0.48, 95% CI: 0.45-0.51) and further to death by 40% (RR=0.60, 95% CI:0.54-0.66). High-risk state patients treated with remdesivir led to a 25% statistically significant reduction in death (RR=0.75, 95% CI: 0.69-0.82). Regarding the outcome of discharge, remdesivir treatment was most effective for medium-risk patients at enrollment (RR: 1.41, 95% CI: 1.35-1.47) followed by high-(RR=1.34, 95% CI: 1.27-1.42) and low-risk patients (RR=1.28, 95% CI: 1.25-1.31).

**Conclusion:** Our results with a simulated two-arm controlled study have provided a new insight into the precision treatment of remdesivir for COVID-19 patients based on risk-stratified efficacy.

## INTRODUCTION

As of May 13^th^, 297,491 deaths out of 4,364,172 confirmed COVID-19 cases, amounting to 7% case-fatality, have been noted during COVID-19 pandemic. Besides, the recovery rate is also less than 30%.^1^ To reduce its death and the length of stay (LOS) in hospitalization resulting from COVID-19, the administration of anti-viral therapy may provide a solution to treat these emerging infected cases.^2-6^ The recent one-arm before-and-after study on the compassionate use of remdesivir for patients with COVID-19 has shown 68% clinical improvement.^7^ Two recent two-arm randomized controlled trials have been conducted to assess its efficacy.^8,9^ One has shown promising but non-significant results on clinical improvement due to insufficient power^8^ whereas the other has shown significant results of the recovery but non-significant results on death from COVID-19 were revealed in the interim report.^9^ We previously demonstrated the efficacy of reducing the risk of death and high-risk state and enhancing the chance of discharge as a result of remedesivir.^10^ Such inconsistent results may be also attributed to the failure to consider risk-stratified efficacy of remedesivir, allowing for the severity of COVID-19, namely, precision treatment for COVID-19. To tackle this issue, the best way is to conduct a stratified randomized controlled trial or adaptive design after interim report to allow for the severity of COVID-19 patients. In order to be timely to reduce the sequelae of COVID-19 pandemic during global emergent crisis, the alternative is to elucidate dynamic of COVID-19 with the exhibition of transition from low-risk state to high-risk state defined by clinical events (e.g. invasive mechanical ventilation and extracorporeal membrane oxygenation (ECMO)) and final to death after the stratification of different risk-state patients at enrollment between the remdesivir-treated and the control group. However, so doing requires a two-arm controlled study.

While waiting for the results of the two-arm randomized controlled trial^9^ or perhaps designing another powerful trial for validating the efficacy of remdesivir, the recent one-arm study published by Grein et al.^7^ provides available empirical individual data to simulate a two-arm controlled study with a modelling approach whereby their data offer an opportunity to elucidate the dynamic of COVID-19 stratified by risk state at enrollment so as to assess the risk-stratified efficacy of remdesivir in reducing the subsequent progression to high-risk state or to final death from COVID-19 and the length of stay of hospitalization. The estimated results derived from such a simulated study enable one to prepare a well-deigned stratified randomized controlled trial with adequate statistical power.

## MATERIAL and METHODS

### Data source

The empirical data used for building up the simulated two-arm controlled study and the following analysis on the efficacy of remdesivir were derived from a recent study on compassionate use of remdesivir for 53 patients with COVID-19 as shown in the Figure 2 of the original article that gives a clear profile of dynamics of disease transition according to the severity of disease from the date of being administered by remdesivir until discharge, death, or the end of study.^7^ The risk state of these patients can be classified into low-(no and low-flow oxygen supplement), medium-(high-flow oxygen supplement and noninvasive positive pressure ventilation), and high-(ECMO and invasive mechanical ventilator) risk states. The aggregated data on each transition mode are listed in **Table 1** after the re-arrangement of the data provided in the original published article.^7^

**Table 1.** Empirical data on all the transition modes of 53 COVID-19 patients by two treatment groups.

| Transition mode |  | Remdesivir |  | Standard care<br>(Control) |  |
| --- | --- | --- | --- | --- | --- |
| From | To | Frequency | % | Frequency | % |
| **On enrollment** |  |  |  |  |  |
|  | **Low** | 12 | (22.6) | 12 | (22.6) |
|  | **Medium** | 7 | (13.2) | 7 | (13.2) |
|  | **High** | 34 | (64.2) | 34 | (64.2) |
| **During follow-up** |  |  |  |  |  |
| **Low** | **Low** | 197 | (87.9) | 197 | (93.4) |
|  | **Medium** | 4 | (1.8) | 4 | (1.9) |
|  | **Discharge** | 23 | (10.3) | 10 | (4.7) |
| **Medium** | **Low** | 12 | (9.1) | - | - |
|  | **Medium** | 113 | (85.6) | 113 | (89.7) |
|  | **High** | 5 | (3.8) | 5 | (4.0) |
|  | **Discharge** | 1 | (0.8) | 7 | (5.6) |
|  | **Death** | 1 | (0.8) | 1 | (0.8) |
| **High** | **Low** | 11 | (2.5) | - | - |
|  | **Medium** | 11 | (2.5) | - | - |
|  | **High** | 403 | (93.3) | 403 | (96.6) |
|  | **Discharge** | 1 | (0.2) | 8 | (1.9) |
|  | **Death** | 6 | (1.4) | 6 | (1.4) |

### Disease transition models for the evolution of COVID-19

Based on data listed in **Table 1**, we simulated a two-arm controlled study following the two transition modes used previously^10^ for patients receiving remdesivir treatment (progressive and regressive mode; low-risk ⇄ medium-risk ⇄ high-risk transitions plus two final outcomes, discharge from each risk state and death after high-risk state) and standard care (only progressive mode; low-risk ➔ medium-risk ➔ high-risk transitions plus two final outcomes, discharge from each risk state and death after high-risk state). For the standard care group, the majority of COVID-19 patients had recovery after discharge from three states, low, medium, and high risk. A proportion of COVID-19 patients deteriorate with risk states and may further progress till death instead of discharge from these risk states. For the remdesivir-treated group, the benefit of clinical improvement is derived from the regression of COVID-19 risk states. Its transition model not only models forward progression but also allows for backward regression from medium-to low-risk and from high-to medium-risk attributed to the use of remdesivir.

### Statistical Analysis

Two continuous-time five state Markov models^11^ were applied to depicting the daily rates of transition between risk states, discharge, and death regarding the disease evolution of COVID-19 patients with remdesivir and standard care following the two transition modes as indicated above. Bayesian Markov Chain Monte Carlo (MCMC) simulation was used to estimate these daily transition rates in the light of likelihood functions based on aggregated data on the remdesivir-treated and the standard care group abstracted form the original article as shown in **Table 1**.

The dynamic curve depicting the evolution of COVID-19 across three risk states and two destinations of discharge and death in one months was derived by using the transition probability matrix for the five defined states given the estimated results on daily transitions rates. The efficacy of remdesivir treatment in accelerating discharge and decelerating subsequent deaths was then assessed by using the two-arm controlled study for a randomized controlled trial equivalent.

Based on the risk-stratified efficacy of remdesivir treatment, we estimated the sample size required for a stratified randomized controlled trial. Given a 5% alpha level and the effect sizes estimated from the simulated two-arm controlled trial, we calculated the sample size for low-, medium-, and high-risk COVID-19 patients required to reach 80% statistical power. The simulated data based on the estimated daily transition rates for the two treatment groups were also generated by using Bayesian Markov Chain Monte Carlo (MCMC) method and are listed in **Table 2** following a stratified randomized controlled design.

**Table 2.** Simulated data on the transition of 416 COVID-19 patients for a stratified randomized controlled trail design.

| Transition mode |  | Remdesivir |  | Standard care |  |
| --- | --- | --- | --- | --- | --- |
| From | To | Frequency | % | Frequency | % |
| **On enrollment** |  |  |  |  |  |
|  | **Low** | 81 | 38.9 | 81 | 38.9 |
|  | **Medium** | 56 | 26.9 | 56 | 26.9 |
|  | **High** | 71 | 34.1 | 71 | 34.1 |
| **During follow-up** |  |  |  |  |  |
| **Low** | **Low** | 618 | 79.7 | 482 | 85.9 |
|  | **Medium** | 44 | 5.7 | 42 | 7.5 |
|  | **High** | 2 | 0.3 | 2 | 0.4 |
|  | **Discharge** | 111 | 14.3 | 35 | 6.2 |
| **Medium** | **Low** | 80 | 14.2 | - | - |
|  | **Medium** | 401 | 71.4 | 465 | 82.7 |
|  | **High** | 72 | 12.8 | 78 | 13.9 |
|  | **Discharge** | 7 | 1.2 | 19 | 3.4 |
|  | **Death** | 2 | 0.4 | - | - |
| **High** | **Low** | 4 | 0.3 | - | - |
|  | **Medium** | 67 | 4.9 | - | - |
|  | **High** | 1241 | 90.8 | 2236 | 96.5 |
|  | **Discharge** | 31 | 2.3 | 51 | 2.2 |
|  | **Death** | 23 | 1.7 | 30 | 1.3 |

## RESULTS

### Empirical data on COVID-19 dynamic abstracted from published article

**Table 1** shows the total of 53 patients on repeated data featuring the change of risk states of COVID-19 receiving remdesivir.^7^ Among these patients, 34 (64.2%) of them were at high-risk state at the enrollment as demonstrated by the Figure 1 of original article (the Invasive Oxygen-support group). **Table 1** also shows the transition of the 53 subjects across three risk states and the final destination of discharge during the one-month study period. The data used for deriving the rate of COVID-19 evolution in the light of two disease transition models for subjects receiving standard care are summarized in **Table 1**. Following the rationales mentioned above, the observed data with regression of risk states were not included for the estimation under the scenario of COVID-19 patients receiving standard care.

**Figure 1.**
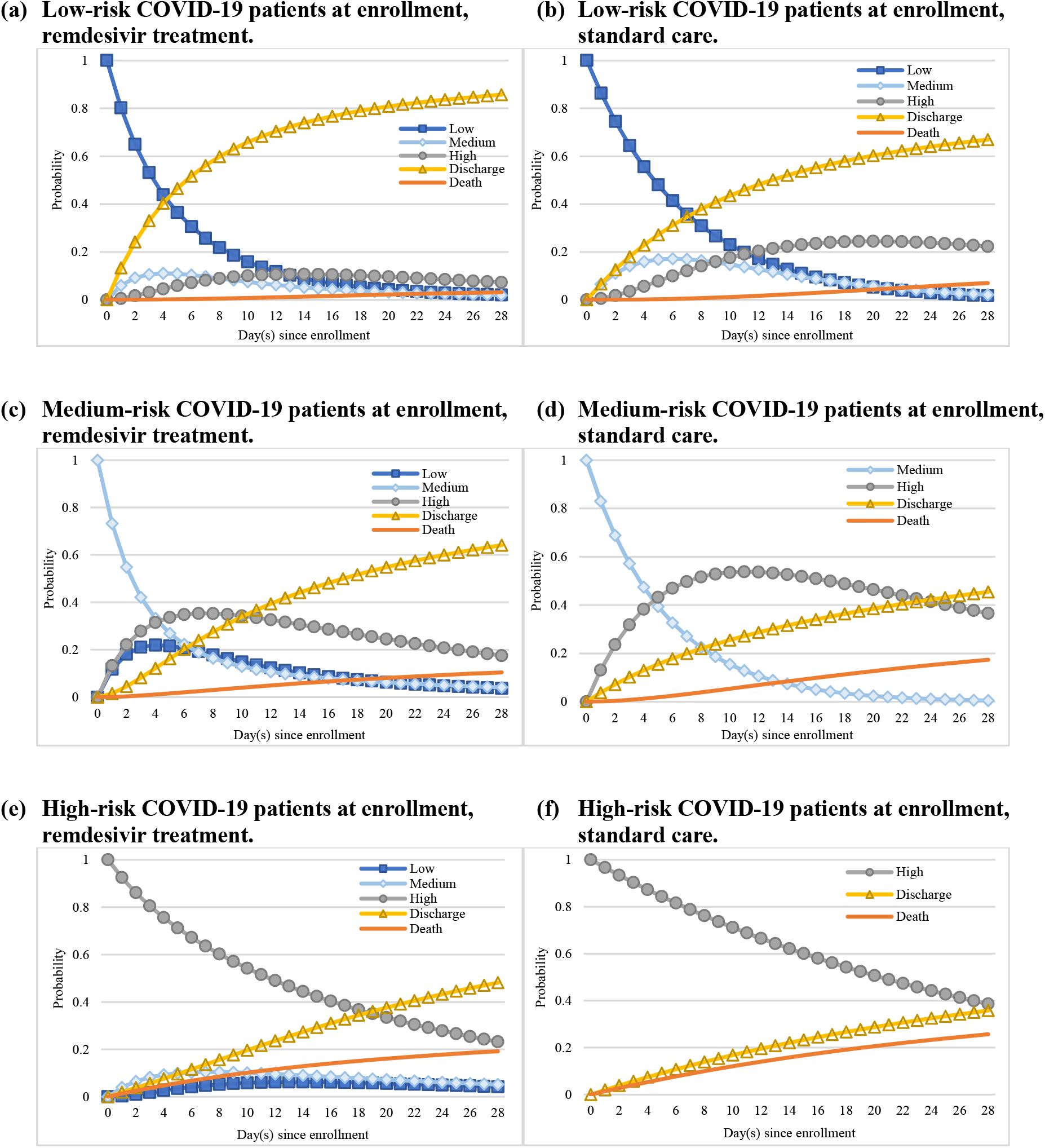
The dynamic of COVID-19 states for remdesivir-treated group (a, c, e) and standard care group (b, d, f) by risk-state at enrollment.

### Daily rates of COVID-19 evolution

**Table 3** shows the estimated results on the daily transition rates for the two continuous-time five-state Markov models. The two daily regression rates were estimated as 0.048 (95% credible interval (CI): 0.027-0.066) from high-risk to medium-risk state and as 0.155 (95% CI: 0.098-0.227) from medium-risk to low-risk state during hospitalization. The higher overall discharge rate regardless of risk states in the treated group as opposed to the control group is mainly explained by the mechanism of two regression rates during hospitalization, resulting in a double proportion of discharge in the low-risk state in the treated group as opposed to the control group (0.149 (95% CI: 0.081-0.206) vs 0.070 (95% CI: 0.030-0.111)). This also accounts for why the rate of discharge in the treated group was lower for the medium-risk state than the control group because a substantial proportion of patients regressing from medium-risk to low-risk state in the treated group during hospitalization. The similar logic is applied to the identical discharge rate between the two groups given the modest regression rate from high-risk to medium-risk state. This simulated treated group shows an adequate fit with the observed data reported in the original article administration of remdesivir (P=0.38, **Table S1** in Supplementary material).

**Table 3.** Estimated results on the daily transition rates between risk states by the two treatment groups

| Transition mode |  | Remdesivir |  | Standard care |  |
| --- | --- | --- | --- | --- | --- |
| From | To | Transition rate | 95% CI | Transition rate | 95% CI |
| **Low** | **Medium** | 0.078 | (0.047, 0.113) | 0.077 | (0.053, 0.106) |
|  | **Discharge** | 0.149 | (0.081, 0.206) | 0.070 | (0.030, 0.111) |
| **Medium** | **Low** | 0.155 | (0.098, 0.227) | - | - - |
|  | **High** | 0.162 | (0.103, 0.217) | 0.147 | (0.098, 0.200) |
|  | **Discharge** | 0.005 | (0.001, 0.016) | 0.040 | (0.006, 0.079) |
| **High** | **Medium** | 0.048 | (0.027, 0.066) | - | - - |
|  | **Discharge** | 0.020 | (0.018, 0.021) | 0.020 | (0.008, 0.034) |
|  | **Death** | 0.014 | (0.005, 0.025) | 0.014 | (0.005, 0.025) |

### The comparisons of dynamic of COVID-19 states by two groups

The benefit of remdesivir treatment was elucidated by the dynamics of COVID-19 across three risk states and the destinations of discharge and death based on the estimated results on daily rates with the application of the two disease transition modes to detailed empirical data. **Figure 1 (a)** shows the daily progression on the dynamic of COVID-19 for the remdesivir-treated and standard care group in patients with low-risk state at enrollment. In line with the estimated results listed in **Table 3**, as the discharge rate for COVID-19 patients were accelerated by remdesivir treatment the probability of discharge (yellow line) was uniformly higher compared with that of the standard care group. For patients progressing to medium- and high-risk, the benefit of regression to a lower risk states resulting from remdesivir was revealed by the uniformly lower probability for medium-(light blue line) and high-risk (gray line) compared with the standard care group (**Figure 1 (b)**). The risk of death (orange line) after one-month follow-up period was also higher in the standard care group compared with the remdesivir-treated group. Similar trends regarding the probabilities of discharge, death, and high-risk state were observed in **Figure 1 (c)** vs **(d)** and **(e)** vs **(f)** showing the dynamic of COVID-19 by two treatment groups for patients with medium- and high-risk state at enrollment.

### Efficacy of remdesivir treatment

Based on the dynamic of COVID-19 evolution elaborated in **Figure 1**, we are able to assess the efficacy of remdesivir treatment compared with standard care at one-month follow-up for low-, medium-, and high-risk COVID-19 patients at enrollment. **Table 4** shows the efficacy of remdesivir for the COIVD-19 patients using the three outcomes of discharge, death, and high-risk state. In all three risk states of COVID-19 patients, the use of remdesivir gave statistically significant higher odds of discharge and lower risks of death and progression to high-risk state. For the remdesivir-treated group, the low-, medium-, and high-risk COVID-19 patients had a higher probability of discharge and lower probability of death and turning into high-risk state compared with the standard care group.

**Table 4.** Estimated results on the one-month probability of risk state, death, and discharge.

| Risk state at enrollment | Transition to states | Remdesivir |  | Standard care |  | RR |  |
| --- | --- | --- | --- | --- | --- | --- | --- |
|  |  | Probability (%) | 95% CI | Probability (%) | 95% CI | Estimate | 95% CI |
| Low | (1) Discharge | 85.7 | (84.8, 86.5) | 66.9 | (65.4, 68.4) | 1.28 | (1.25, 1.31) |
|  | (2) High-risk | 7.3 | (6.8, 7.8) | 22.3 | (21.2, 23.3) | 0.33 | (0.30, 0.35) |
|  | (3) Death | 3.1 | (2.9, 3.4) | 7.0 | (6.6, 7.6) | 0.45 | (0.39, 0.50) |
|  | (2)+(3) | 10.4 | (9.8, 11.1) | 29.3 | (28.0, 30.6) | 0.36 | (0.33, 0.38) |
| Medium | (1) Discharge | 64.0 | (62.8, 65.0) | 45.5 | (43.7, 47.3) | 1.41 | (1.35, 1.47) |
|  | (2) High-risk | 17.6 | (16.9, 18.5) | 36.6 | (35.1, 38.3) | 0.48 | (0.45, 0.51) |
|  | (3) Death | 10.4 | (9.8, 11.2) | 17.4 | (16.2, 18.5) | 0.60 | (0.54, 0.66) |
|  | (2)+(3) | 28.1 | (27.0, 29.1) | 54.0 | (52.1, 55.7) | 0.52 | (0.49, 0.55) |
| High | (1) Discharge | 48.0 | (47.2, 48.8) | 35.7 | (33.7, 37.6) | 1.34 | (1.27, 1.42) |
|  | (2) High-risk | 23.4 | (22.6, 24.3) | 38.6 | (36.6, 40.3) | 0.61 | (0.57, 0.64) |
|  | (3) Death | 19.2 | (18.1, 20.5) | 25.7 | (24.1, 27.3) | 0.75 | (0.69, 0.82) |
|  | (2)+(3) | 42.6 | (41.6, 43.6) | 64.3 | (62.4, 66.3) | 0.66 | (0.64, 0.69) |

The efficacy of remdesivir in accelerating discharge of COVID-19 patients was most prominent for the medium-risk group (relative risk (RR)=1.41, 95 CI: 1.35-1.47, **Table 4**, final column) followed by the high-risk group (RR=1.34, 95% CI: 1.27-1.42) and the low-risk group (RR=1.28, 95% CI: 1.25-1.31).

For low-risk patient at enrollment, **Table 4** shows remdesivir led to the highest efficacy in reducing subsequent progression to high-risk state by 67% (RR=0.33, 95% CI: 0.30-0.35) and to final death by 55% (RR=0.45, 95% CI: 0.39-0.50). For the medium-risk patients, less but still statistically significant efficacy results were noted in reducing progression to high-risk state by 52% (RR=0.48, 95% CI: 0.45-0.51) and further to death by 40% (RR=0.60, 95% CI:0.54-0.66). High-risk state patients treated with remdesivir also led to a 25% statistically significant reduction in death (RR=0.75, 95% CI: 0.69-0.82).

### Sample size determination for designing a stratified randomized controlled trial

Given the effect size mentioned above for the primary outcome combing death and progress to high-risk state for the low-, medium-, and high-risk COVID-19 patient at enrollment, we estimated the sample size required for preparing a stratified randomized controlled clinical trial with adequate statistical power. Assuming a 5% alpha level, 208 COVID-19 patients are required, consisting of 81 low-risk, 56 medium-risk, and 71 high-risk patients for each arm in order to reach 80% statistical power. The summary of the simulated data given the estimated sample size are listed in **Table 2**.

## DISCUSSION

While a substantial proportion of COVID-19 cases have been spawned from a cascade of outbreaks in five continental countries from January until May, the global disease burden has been imposed on enormous deaths from COVID-19 as well as slow recovery after the ascertainment of COVID-19 cases. The slow recovery may increase the possibility of transmission and infectious period so as to spur the next outbreak after contact with the susceptible. To reduce these two burdens, one of solutions is to have the better use of potential promising anti-viral therapy like remdesivir, which has been proposed for compassionated use in total of 53 patients and demonstrated 68% clinical improvement.^7^ Although it is a one-arm study, a simulated two-arm controlled study was therefore conducted to further prove the efficacy of remdesivir in reducing death by 29% (95% CI: 22-35%) and high-risk state of COVID-19 patients by 45% (95% CI: 42-48%).^10^ It is also very effective in halving the median time of length of stay in hospitalization.^10^

As the efficacy of remdesivir may vary with the severity of COVID-19 at enrollment how to provide risk-stratified efficacy evidence plays an important role in precision medicine for the optimal treatment of COVID-19. To achieve this goal, we may need another stratified randomized controlled trail or an adaptive randomized controlled trial. While waiting for these well-designed trials, the alternative is to elucidate dynamics of COVID-19 from low-risk state to final death or recovery based on a simulated two-arm controlled trial, which becomes the main contribution of this study.

By using the two disease transition models together with the empirical data abstracted from the recent study on the compassion use of remdesivir, we estimated the daily transition rates and elucidated the dynamics of COVID-19 between different risk states that provides an insight into the clinical mechanism of COVID-19 disease progression and also regression resulting from remdesivir treatment. The kinetic curves as shown in **Figure 1** clearly depict he evolution of COVID-19 from low-risk to high-risk and also the two primary outcomes, recovery and death for the two treatment groups. These curves provide the basis for the evaluation of the efficacy of remdesivir in decelerating the force of progression and accelerating the force of regression, resulting in the reduction of death and the length of hospitalization due to recovery.

According to our findings on the dynamics of kinetic curves, the most benefit of reducing the risk of death and progression to high-risk state as a result of remdesivir treatment commences with low-risk state patients whereas the major impact on the reduction of time to discharge is attributed to the treatment offered for the medium-risk group. The simulated data on a stratified two-arm controlled study offer an opportunity to determine sample size by the stratification of severity of COVID-19 at enrollment, which would be helpful in the preparation of a well-designed stratified randomized controlled trial.

The additional advantages of our secondary analysis of the empirical data from the original one-arm compassionate study on the use of remdesivir,^7^ while using the simulated two-arm controlled study, are three-fold. Firstly, we improved the weakness of a lacking control group in the original study as a proportion of COVID-19 patients may be discharged with the recovery dispensing with the use of remdesivir. Second, such a simulated two-arm study design enables us to evaluate the efficacy of remdesivir with clear primary endpoints including death and discharge rather than only based on the clinical improvement before and after the use of remdesivir defined by live discharge from hospital, a decrease of at least 2 points from baseline on the supplementary oxygen use, or both as used in the original article.^7^ Third, the results from such a simulated two-arm controlled study are supposed to be closer to those using a real two-arm randomized controlled trial in the near future in terms of intention-to-treat analysis principle.

The results on the efficacy of remdesivir in reducing death and length of stay resulting from COVID-19 have also two significant implications for containing COVID-19 pandemic. Firstly, it reduces the sequelae of COVID-19 and also accelerates its recovery. Besides, the administration of remdesivir may also reduce transmission probability and also infectious time of the infective in contact with the susceptible subjects. Such an efficacy of prophylactic and therapeutic anti-viral therapy has been demonstrated in influenza.^12,13^

Our results were derived from the data on 53 COVID-19 patients receiving compassionate remdesivir treatment,^7^ which may be subject to volunteer bias. This would limit the generalizability of our study to other external populations but this may not be serious during pandemic period. In addition, the results should be validated by a randomized controlled trial, which can be facilitated by our study for an efficient design.

In conclusion, we demonstrated the risk-stratified efficacy of remdesivir for COVID-19 patients by using a simulated two-arm controlled study. Our findings have a significant implication for precision anti-viral therapy administered by COVID-19 patients.

## Data Availability

All data and codes are available from author on request.

## Author contributions

HHC, CYH, and CCL contributed to the study concept and designed. CCL, TYL and WJC contributed to the acquisition of data. AMFY, SLSC, TYL, and WJC contributed to the development of method, statistical analysis, and computer programming. CCL, AMFY, SLSC contributed to the interpretation of results. TYL and WJC drafted the manuscript. HHC, AMFY, SLSC and CYH revised the manuscript critically for intellectual content. All authors gave final approval of the manuscript.

## Acknowledgements

This work was financially supported by the “Innovation and Policy Center for Population Health and Sustainable Environment (Population Health Research Center, PHRC), College of Public Health, National Taiwan University” from The Featured Areas Research Center Program within the framework of the Higher Education Sprout Project by the Ministry of Education (MOE) in Taiwan.

## Data and materials availability

All data and codes are available from author on request.

## Supplementary material

**Table S1.** Model fitting statistics for observed and expected risk states.

| **COVID-19**<br>**Status** | **Frequency** |  |
| --- | --- | --- |
|  | **Observed** | **Expected** |
| **Low** | 232 | 213.8 |
| **Medium** | 135 | 138.7 |
| **High** | 442 | 447.4 |
| **Discharge** | 25 | 34.3 |
| **Death** | 7 | 6.9 |

\* Chi-squared value=4.23, $P=0.38$

